# Machine Learning Applications and Advancements in Alcohol Use Disorder: A Systematic Review

**DOI:** 10.1101/2022.06.06.22276057

**Authors:** Myrna Hurtado, Anna Siefkas, Misty M Attwood, Zohora Iqbal, Jana Hoffman

## Abstract

**Background:** Alcohol use disorder (AUD) is a chronic mental disorder that leads to harmful, compulsive drinking patterns that can have serious consequences. Advancements are needed to overcome current barriers in diagnosis and treatment of AUD.

**Objectives:** This comprehensive review analyzes research efforts that apply machine learning (ML) methods for AUD prediction, diagnosis, treatment and health outcomes.

**Methods:** A systematic literature review was conducted. A search performed on 12/02/2020 for published articles indexed in Embase and PubMed Central with AUD and ML-related terms retrieved 1,628 articles. We identified those that used ML-based techniques to diagnose AUD or make predictions concerning AUD or AUD-related outcomes. Studies were excluded if they were animal research, did not diagnose or make predictions for AUD or AUD-related outcomes, were published in a non-English language, only used conventional statistical methods, or were not a research article.

**Results:** After full screening, 70 articles were included in our review. Algorithms developed for AUD predictions utilize a wide variety of different data sources including electronic health records, genetic information, neuroimaging, social media, and psychometric data. Sixty-six of the included studies displayed a high or moderate risk of bias, largely due to a lack of external validation in algorithm development and missing data.

**Conclusions:** There is strong evidence that ML-based methods have the potential for accurate predictions for AUD, due to the ability to model relationships between variables and reveal trends in data. The application of ML may help address current underdiagnosis of AUD and support those in recovery for AUD.

## INTRODUCTION

Alcohol misuse is characterized by unhealthy drinking patterns, such as binge drinking and heavy alcohol use, that increases the risk of alcohol use disorder (AUD). (1) AUD is strongly associated with morbidity and mortality, and a major public health burden globally.(2) One study reported 93,296 deaths per year in the United States (U.S.) due to excessive alcohol consumption, with an average of 29 years of life lost per early death.(3) More than 99 million disability-adjusted life-years were attributable to alcohol use in 2016,(2) and alcohol misuse is considered to be one of the leading causes of preventable deaths in the U.S.(4) Two main challenges associated with combating AUD are underdiagnosis and unsuccessful treatment outcomes. Although the U.S. Preventive Services Task Force has recommended screening in primary care settings to identify and curtail unhealthy alcohol use,(5) alcohol misuse screening rates remain low.(6) Data from the Behavioral Risk Factor Surveillance System revealed that low rates arise predominantly from missed screening opportunities during primary care visits rather than inadequate access to healthcare.(7) Relapse is quite common among individuals with AUD; 40% to 60% of patients relapse within the first year after treatment completion.(8) Diagnosis of, and accurate predictions for, individuals with AUD are crucial for successful treatment outcomes as well as prevention of other resultant morbidities.

The application of artificial intelligence, and specifically machine learning (ML) in healthcare, have the potential to revolutionize approaches in medicine.(9) ML has been leveraged to improve disease prediction and detection, medical imaging, drug discovery and development, genetic analysis, treatment courses, and outcomes predictions.(9,10) ML in healthcare settings may be used to support clinicians in the decision-making process by providing accurate, timely, unbiased, and convenient access to data and analysis. ML methods may also be applied in AUD research. We have previously developed ML for the accurate and early prediction of septic shock(11), and mortality(12) for the AUD population in the intensive care unit. The purpose of this systematic review is to evaluate the use of ML to enhance current diagnostic and outcome prediction approaches for individuals with alcohol misuse and AUD. Definitions of AUD-related terms(13) and ML can be found in Box 1.

### Box 1

Definitions of alcohol use disorder-related terms.

#### Alcohol consumption

##### Alcohol use disorder (AUD)

A brain disorder that results in compulsive drinking despite negative consequences on social life, employment and health. Also referred to as alcoholism, alcohol dependence, addiction or abuse.

##### Binge drinking

Drinking patterns that result in a blood alcohol concentration (BAC) of 0.08% or higher.

##### Heavy alcohol use

Drinking more than 3 drinks in any given day or exceeding more than 7 drinks a week for women or more than 4 drinks in any given day or more than 14 drinks a week for men.

##### Machine learning

An artificial intelligence technique to develop computer algorithms that analyze and learn from patterns in prior data to predict outcomes.

## METHODS

Systematic searches for studies of machine learning applications in AUD in the electronic databases PubMed Central and Embase were conducted by AS on December 2nd, 2020 in accordance with Preferred Reporting Items for Systematic Reviews and Meta-Analyses (PRISMA) guidelines.(14) The search parameters included all studies published prior to the search date and included the search terms “alcohol related disorder” and relevant synonyms coupled with “machine learning” and relevant synonyms (Supplementary Table 1).

### Search strategy and selection criteria

Search results were collected in Google Sheets, and duplicates were first removed in the Zotero reference manager (Corporation for Digital Scholarship, VA, U.S.). Title and abstract screening were conducted by 4 individuals: AS, AGS, NZ, and ZI, where each entry was independently screened by two reviewers. All disagreements were screened by a third reviewer whose input served as a tiebreaker. Full text screening was then conducted by MH, MMA, AGS, and AS, where each manuscript was screened by two reviewers independently, and disagreements were again screened by a third reviewer. For each study, the following information was collected by ZI, DE, MH, and MMA: study design, study aim, results, and clinical impact.

Studies were included if the aims included the following topics: (i) alcohol withdrawal; (ii) genetics or genome-wide association study (GWAS); (iii) diagnosis, treatment, prediction of AUD as the primary disorder; (iv) prediction of AUD treatment-seeking behavior, recovery, or treatment outcome; (v) AUD prediction or identification using experimental data; and (vi) alcohol and drug use. Studies were excluded if they were animal research, did not examine AUD as the primary disorder but rather examined related disease states where AUD was a risk phenotype/predictor, did not use ML or only used conventional statistical methods, published in a non-English language, were presented in conference abstracts and poster presentations, or were review articles, meta-analyses, opinions, or editorials, or if full-text for the study could not be found. ML generates predictive patterns from relationships between variables, while conventional statistical methods draw inferences. (Figure 1)

**Figure 1.**
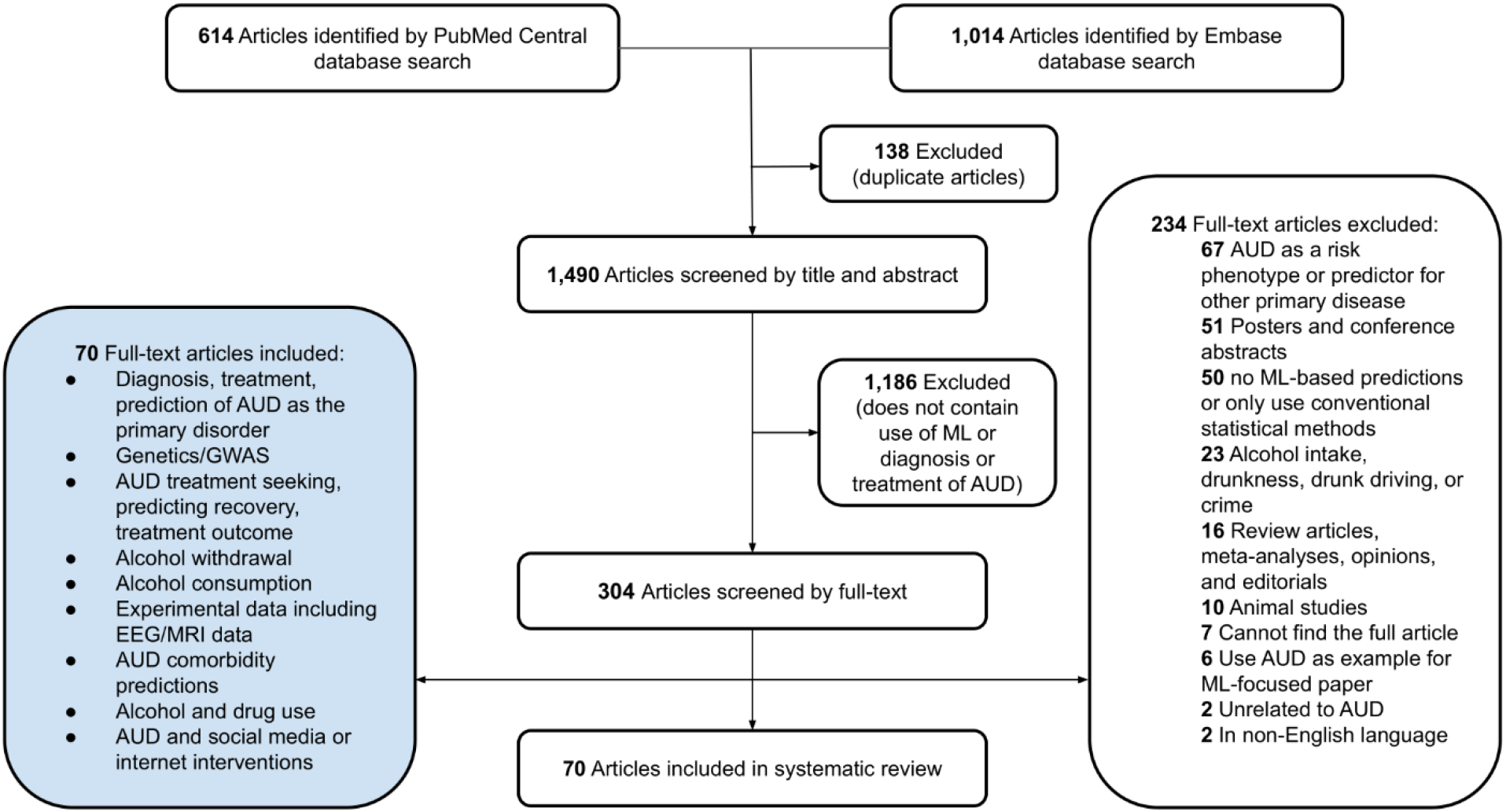
Flow diagram of the article selection process following PRISMA guidelines. Abbreviations: Alcohol use disorder (AUD); electroencephalography (EEG); machine learning (ML); magnetic resonance imaging (MRI).

### Risk of bias assessment

Risk of bias was assessed based on the following criteria: source of data, missing data in sample, lack of external validation, or other apparent sources of risk of bias. For each of these criteria, the reviewer scored the study as “low”, “moderate” or “high” based on the possibility of bias; details on how the risk of bias and overall score was determined may be found in Supplementary Table 2.

## RESULTS

A total of 1,628 studies were extracted from our search and after the removal of 138 duplicate articles, 1,490 were screened by title and abstract (see Figure 1 for article screening process). Following screening, 1,186 articles were excluded because they did not pertain to the diagnosis or treatment of AUD or mention the use of ML. The remaining 304 articles were screened by full-text reviews. A total of 70 studies met our inclusion criteria and were included in our review. The summarized study aims and clinical impacts are presented in Table 1 and Table 2. Additional study characteristics such as type of ML algorithm, sample size, type of data, ethnicity/race, age, gender, comparator and/or competitor can be found in Supplementary Table 3. The included studies apply a variety of different ML techniques and utilize electronic health records (EHR), genetic, neuroimaging, psychometric, and internet-based data to make predictions with regard to AUD and AUD-related outcomes (Figure 2).

**Table 1.**
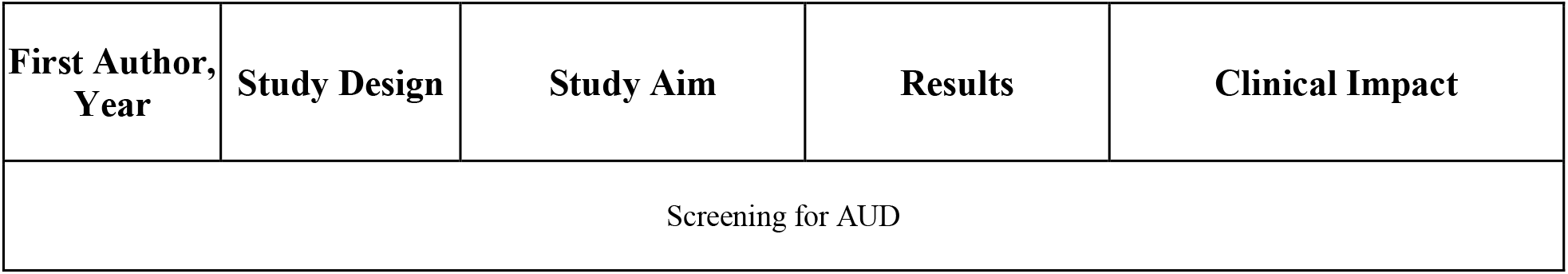
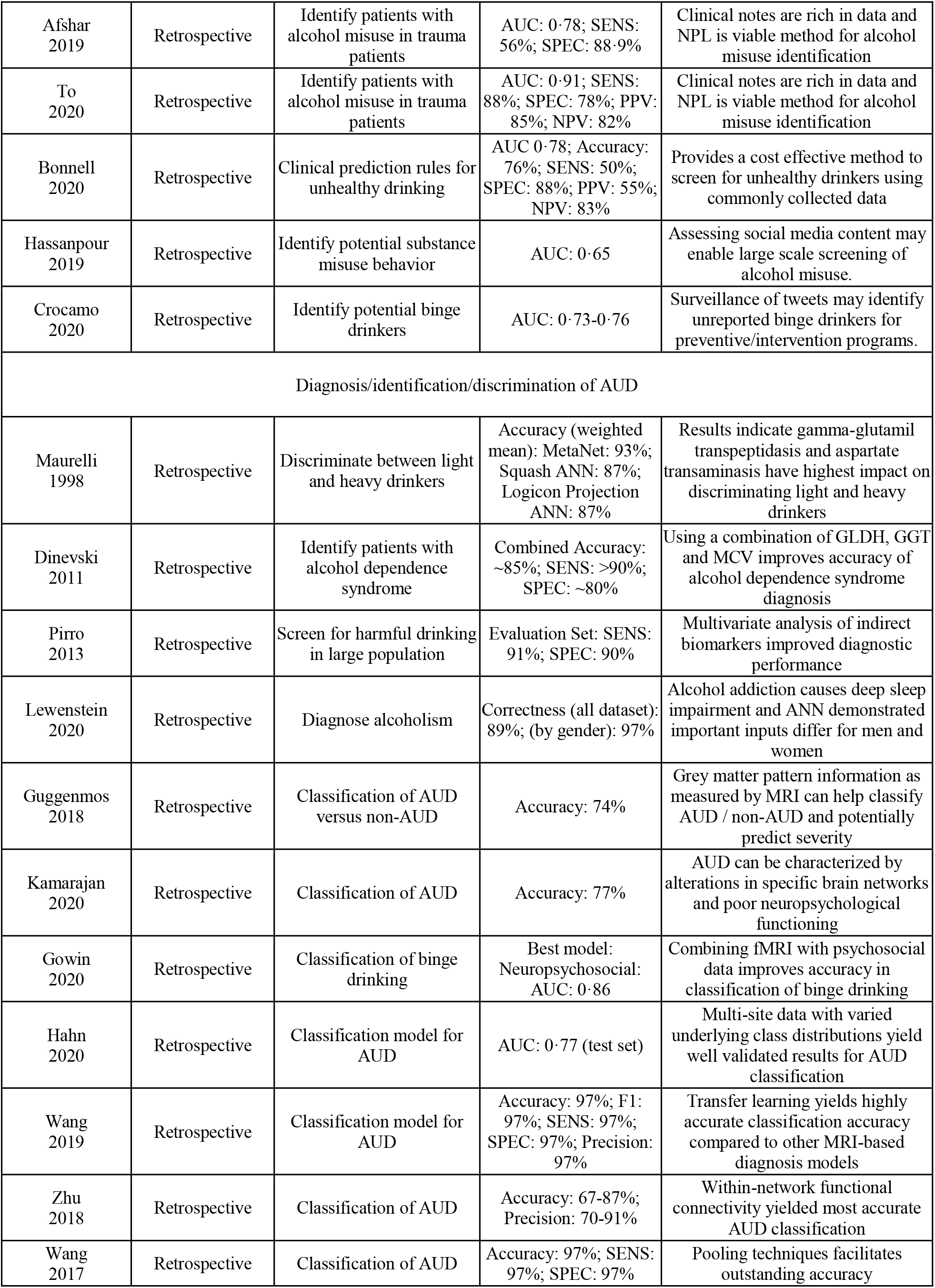
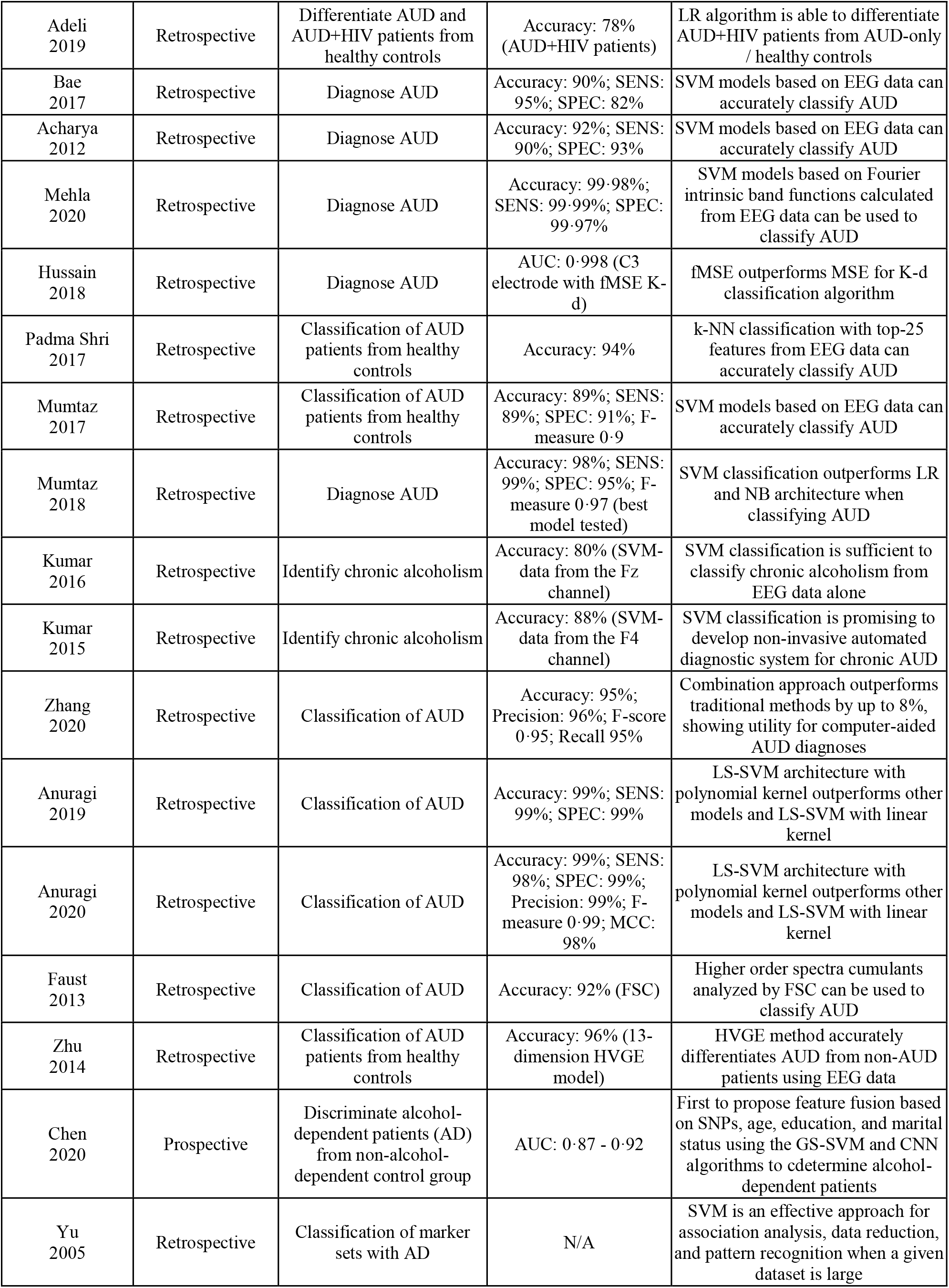
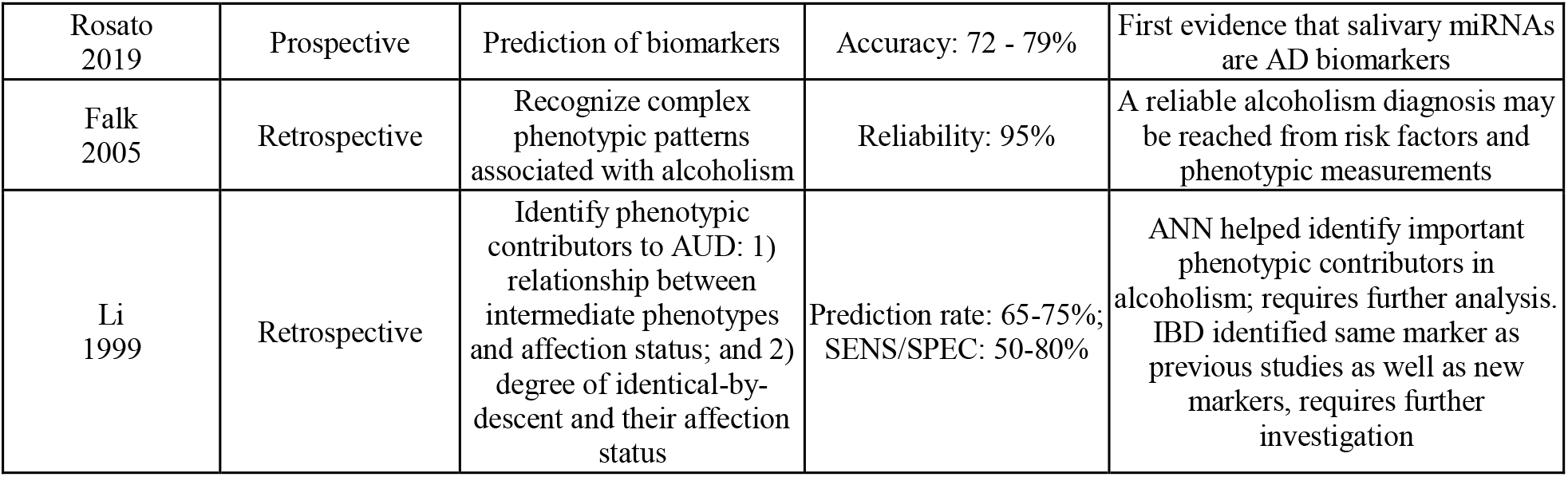
Summary of reviewed articles pertaining to screening for AUD and diagnosis/identification/discrimination of AUD.

| First Author, Year | Study Design | Study Aim | Results | Clinical Impact |
| --- | --- | --- | --- | --- |
| Screening for AUD |  |  |  |  |
| Afshar 2019 | Retrospective | Identify patients with alcohol misuse in trauma patients | AUC: 0.78; SENS: 56%; SPEC: 88.9% | Clinical notes are rich in data and NPL is viable method for alcohol misuse identification |
| To 2020 | Retrospective | Identify patients with alcohol misuse in trauma patients | AUC: 0.91; SENS: 88%; SPEC: 78%; PPV: 85%; NPV: 82% | Clinical notes are rich in data and NPL is viable method for alcohol misuse identification |
| Bonnell 2020 | Retrospective | Clinical prediction rules for unhealthy drinking | AUC 0.78; Accuracy: 76%; SENS: 50%; SPEC: 88%; PPV: 55%; NPV: 83% | Provides a cost effective method to screen for unhealthy drinkers using commonly collected data |
| Hassanpour 2019 | Retrospective | Identify potential substance misuse behavior | AUC: 0.65 | Assessing social media content may enable large scale screening of alcohol misuse. |
| Crocamo 2020 | Retrospective | Identify potential binge drinkers | AUC: 0.73-0.76 | Surveillance of tweets may identify unreported binge drinkers for preventive/intervention programs. |
| Diagnosis/identification/discrimination of AUD |  |  |  |  |
| Maurelli 1998 | Retrospective | Discriminate between light and heavy drinkers | Accuracy (weighted mean): MetaNet: 93%; Squash ANN: 87%; Logicon Projection ANN: 87% | Results indicate gamma-glutamyl transpeptidase and aspartate transaminase have highest impact on discriminating light and heavy drinkers |
| Dinevski 2011 | Retrospective | Identify patients with alcohol dependence syndrome | Combined Accuracy: ~85%; SENS: >90%; SPEC: ~80% | Using a combination of GDLH, GGT and MCV improves accuracy of alcohol dependence syndrome diagnosis |
| Pirro 2013 | Retrospective | Screen for harmful drinking in large population | Evaluation Set: SENS: 91%; SPEC: 90% | Multivariate analysis of indirect biomarkers improved diagnostic performance |
| Lewenstein 2020 | Retrospective | Diagnose alcoholism | Correctness (all dataset): 89%; (by gender): 97% | Alcohol addiction causes deep sleep impairment and ANN demonstrated important inputs differ for men and women |
| Guggenmos 2018 | Retrospective | Classification of AUD versus non-AUD | Accuracy: 74% | Grey matter pattern information as measured by MRI can help classify AUD / non-AUD and potentially predict severity |
| Kamarajan 2020 | Retrospective | Classification of AUD | Accuracy: 77% | AUD can be characterized by alterations in specific brain networks and poor neuropsychological functioning |
| Gowin 2020 | Retrospective | Classification of binge drinking | Best model: Neuropsychosocial: AUC: 0.86 | Combining fMRI with psychosocial data improves accuracy in classification of binge drinking |
| Hahn 2020 | Retrospective | Classification model for AUD | AUC: 0.77 (test set) | Multi-site data with varied underlying class distributions yield well validated results for AUD classification |
| Wang 2019 | Retrospective | Classification model for AUD | Accuracy: 97%; F1: 97%; SENS: 97%; SPEC: 97%; Precision: 97% | Transfer learning yields highly accurate classification accuracy compared to other MRI-based diagnosis models |
| Zhu 2018 | Retrospective | Classification of AUD | Accuracy: 67-87%; Precision: 70-91% | Within-network functional connectivity yielded most accurate AUD classification |
| Wang 2017 | Retrospective | Classification of AUD | Accuracy: 97%; SENS: 97%; SPEC: 97% | Pooling techniques facilitates outstanding accuracy |
| Adeli<br>2019 | Retrospective | Differentiate AUD and AUD+HIV patients from healthy controls | Accuracy: 78% (AUD+HIV patients) | LR algorithm is able to differentiate AUD+HIV patients from AUD-only / healthy controls |
| Bae<br>2017 | Retrospective | Diagnose AUD | Accuracy: 90%; SENS: 95%; SPEC: 82% | SVM models based on EEG data can accurately classify AUD |
| Acharya<br>2012 | Retrospective | Diagnose AUD | Accuracy: 92%; SENS: 90%; SPEC: 93% | SVM models based on EEG data can accurately classify AUD |
| Mehla<br>2020 | Retrospective | Diagnose AUD | Accuracy: 99.98%; SENS: 99.99%; SPEC: 99.97% | SVM models based on Fourier intrinsic band functions calculated from EEG data can be used to classify AUD |
| Hussain<br>2018 | Retrospective | Diagnose AUD | AUC: 0.998 (C3 electrode with fMSE K-d) | fMSE outperforms MSE for K-d classification algorithm |
| Padma Shri<br>2017 | Retrospective | Classification of AUD patients from healthy controls | Accuracy: 94% | k-NN classification with top-25 features from EEG data can accurately classify AUD |
| Mumtaz<br>2017 | Retrospective | Classification of AUD patients from healthy controls | Accuracy: 89%; SENS: 89%; SPEC: 91%; F-measure 0.9 | SVM models based on EEG data can accurately classify AUD |
| Mumtaz<br>2018 | Retrospective | Diagnose AUD | Accuracy: 98%; SENS: 99%; SPEC: 95%; F-measure 0.97 (best model tested) | SVM classification outperforms LR and NB architecture when classifying AUD |
| Kumar<br>2016 | Retrospective | Identify chronic alcoholism | Accuracy: 80% (SVM-data from the Fz channel) | SVM classification is sufficient to classify chronic alcoholism from EEG data alone |
| Kumar<br>2015 | Retrospective | Identify chronic alcoholism | Accuracy: 88% (SVM-data from the F4 channel) | SVM classification is promising to develop non-invasive automated diagnostic system for chronic AUD |
| Zhang<br>2020 | Retrospective | Classification of AUD | Accuracy: 95%; Precision: 96%; F-score 0.95; Recall 95% | Combination approach outperforms traditional methods by up to 8%, showing utility for computer-aided AUD diagnoses |
| Anuragi<br>2019 | Retrospective | Classification of AUD | Accuracy: 99%; SENS: 99%; SPEC: 99% | LS-SVM architecture with polynomial kernel outperforms other models and LS-SVM with linear kernel |
| Anuragi<br>2020 | Retrospective | Classification of AUD | Accuracy: 99%; SENS: 98%; SPEC: 99%; Precision: 99%; F-measure 0.99; MCC: 98% | LS-SVM architecture with polynomial kernel outperforms other models and LS-SVM with linear kernel |
| Faust<br>2013 | Retrospective | Classification of AUD | Accuracy: 92% (FSC) | Higher order spectra cumulants analyzed by FSC can be used to classify AUD |
| Zhu<br>2014 | Retrospective | Classification of AUD patients from healthy controls | Accuracy: 96% (13-dimension HVGE model) | HVGE method accurately differentiates AUD from non-AUD patients using EEG data |
| Chen<br>2020 | Prospective | Discriminate alcohol-dependent patients (AD) from non-alcohol-dependent control group | AUC: 0.87 - 0.92 | First to propose feature fusion based on SNPs, age, education, and marital status using the GS-SVM and CNN algorithms to determine alcohol-dependent patients |
| Yu<br>2005 | Retrospective | Classification of marker sets with AD | N/A | SVM is an effective approach for association analysis, data reduction, and pattern recognition when a given dataset is large |
| Rosato<br>2019 | Prospective | Prediction of biomarkers | Accuracy: 72 - 79% | First evidence that salivary miRNAs are AD biomarkers |
| Falk<br>2005 | Retrospective | Recognize complex phenotypic patterns associated with alcoholism | Reliability: 95% | A reliable alcoholism diagnosis may be reached from risk factors and phenotypic measurements |
| Li<br>1999 | Retrospective | Identify phenotypic contributors to AUD: 1) relationship between intermediate phenotypes and affection status; and 2) degree of identical-by-descent and their affection status | Prediction rate: 65-75%; SENS/SPEC: 50-80% | ANN helped identify important phenotypic contributors in alcoholism; requires further analysis. IBD identified same marker as previous studies as well as new markers, requires further investigation |

**Table 2.**
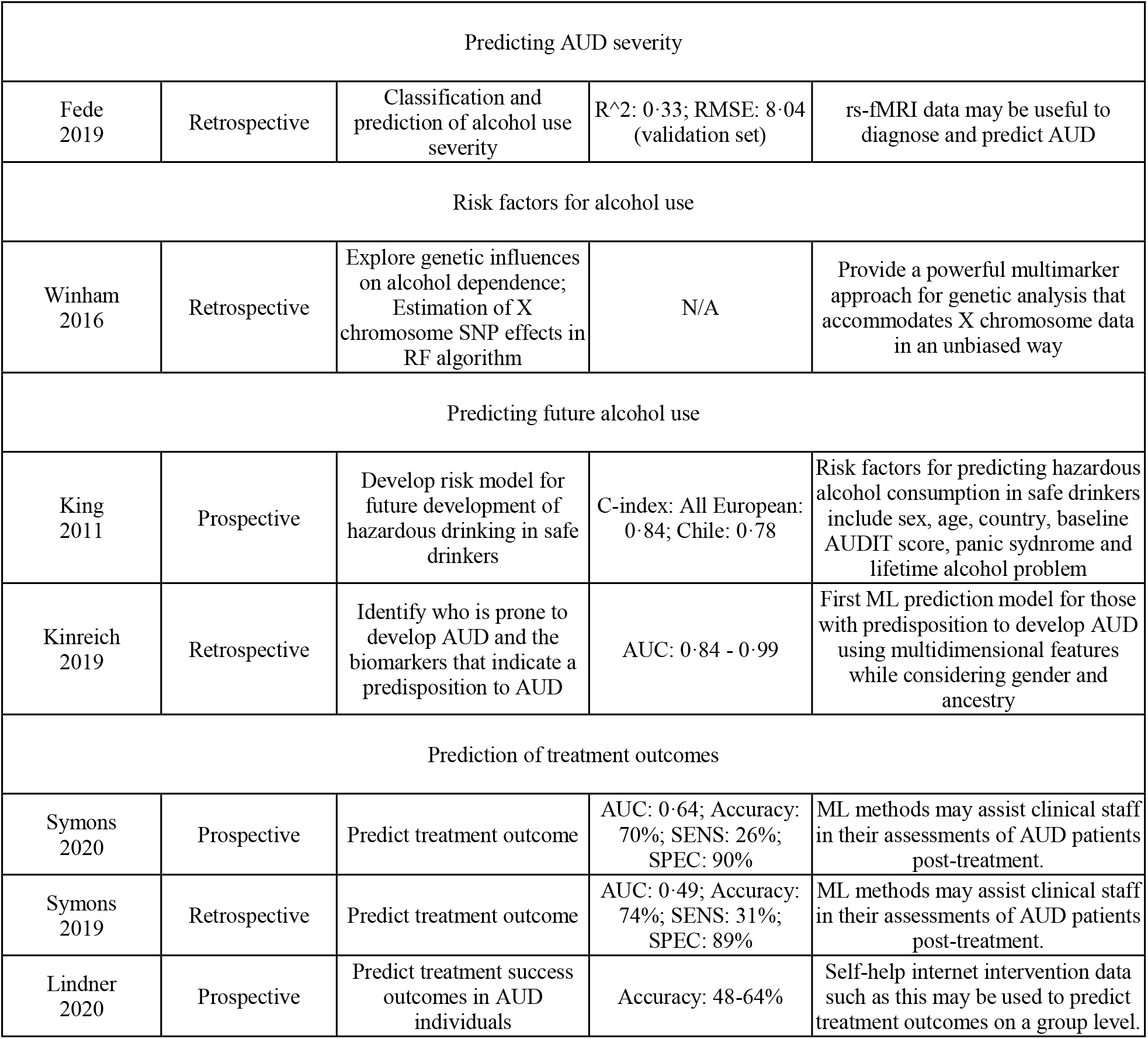
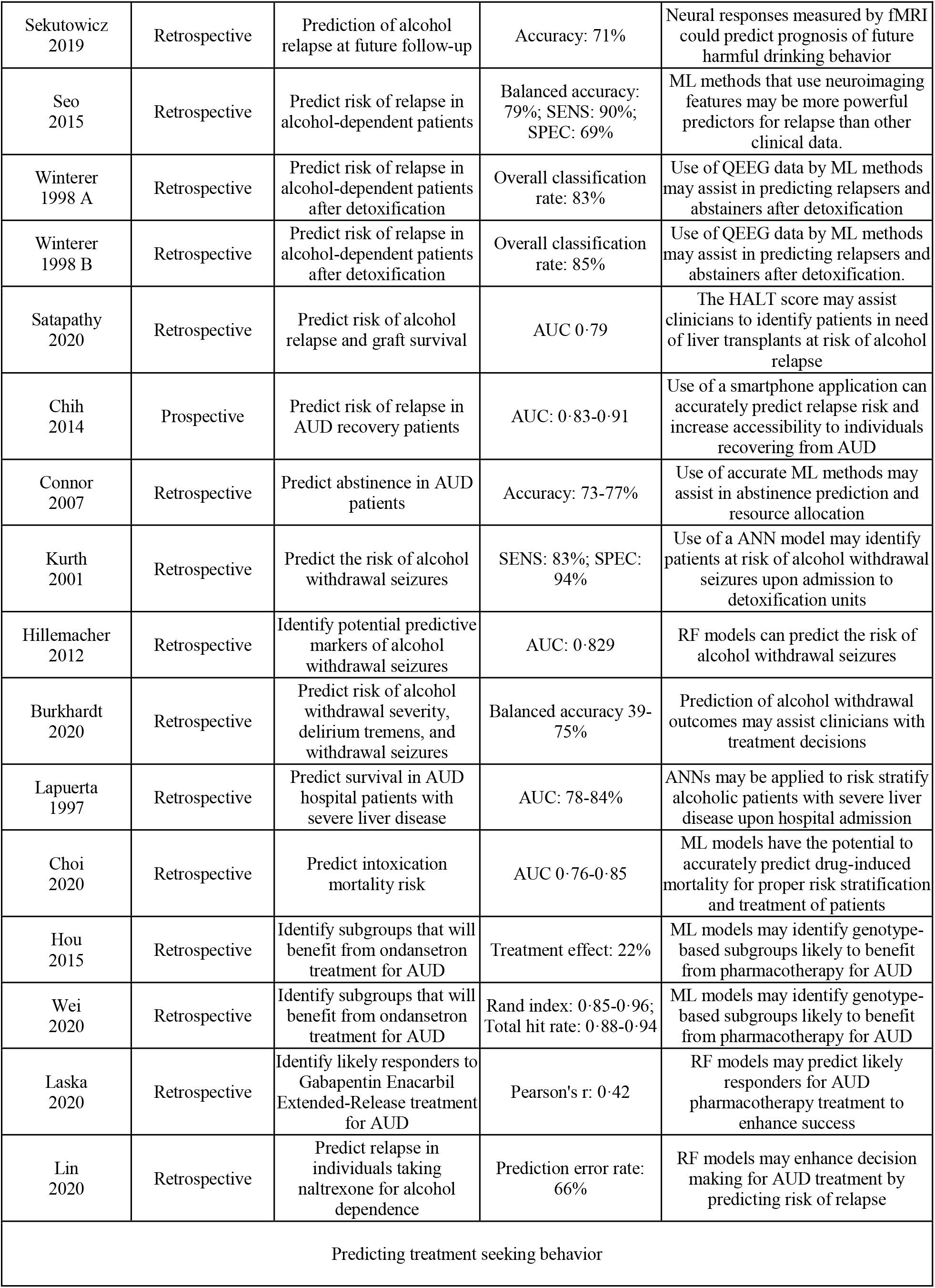
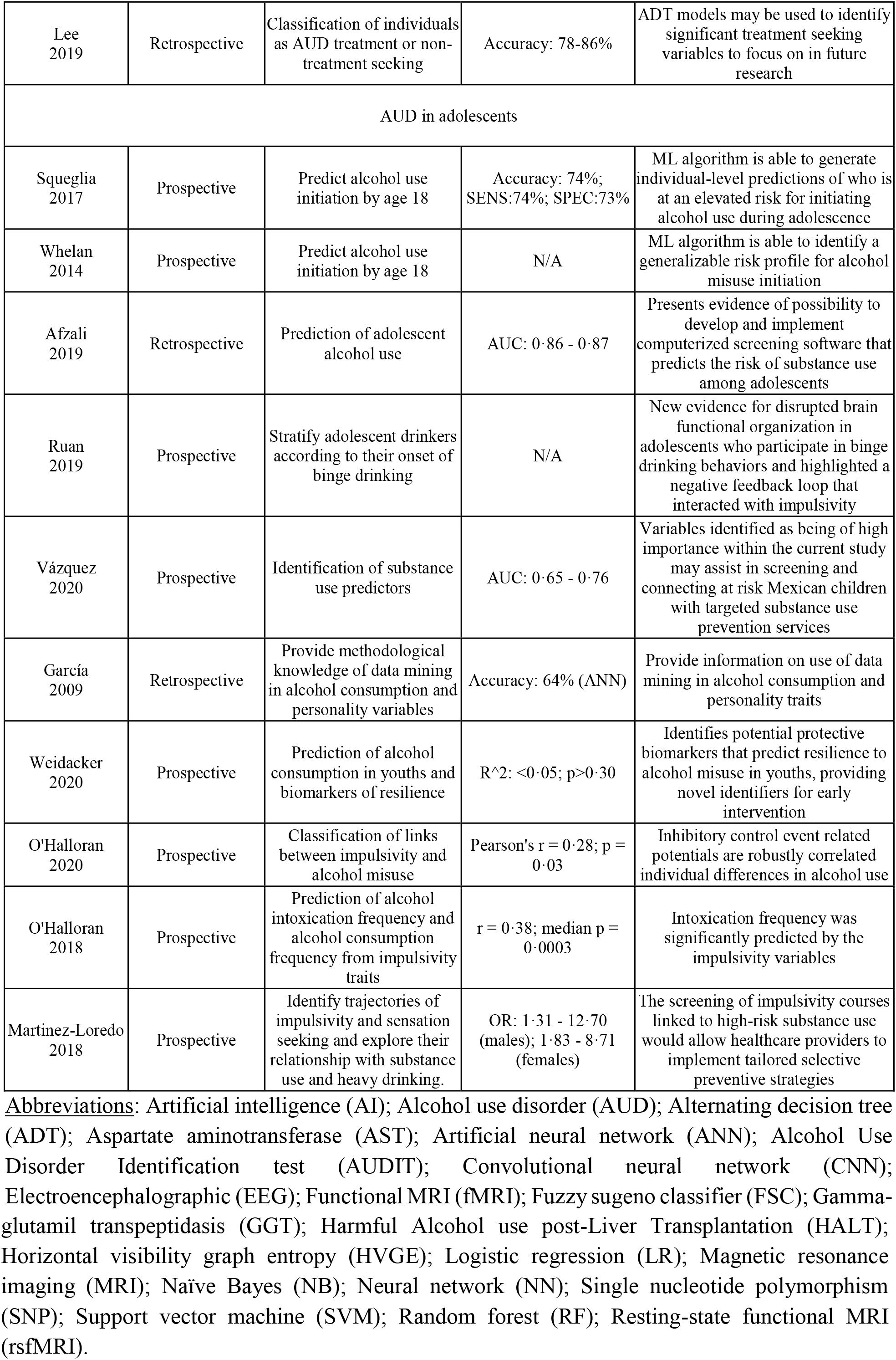
Summary of reviewed articles pertaining to: predicting AUD severity; risk factors for alcohol use; predicting future alcohol use; prediction of treatment outcomes; predicting treatment seeking behavior; and AUD in adolescents.

**Figure 2.**
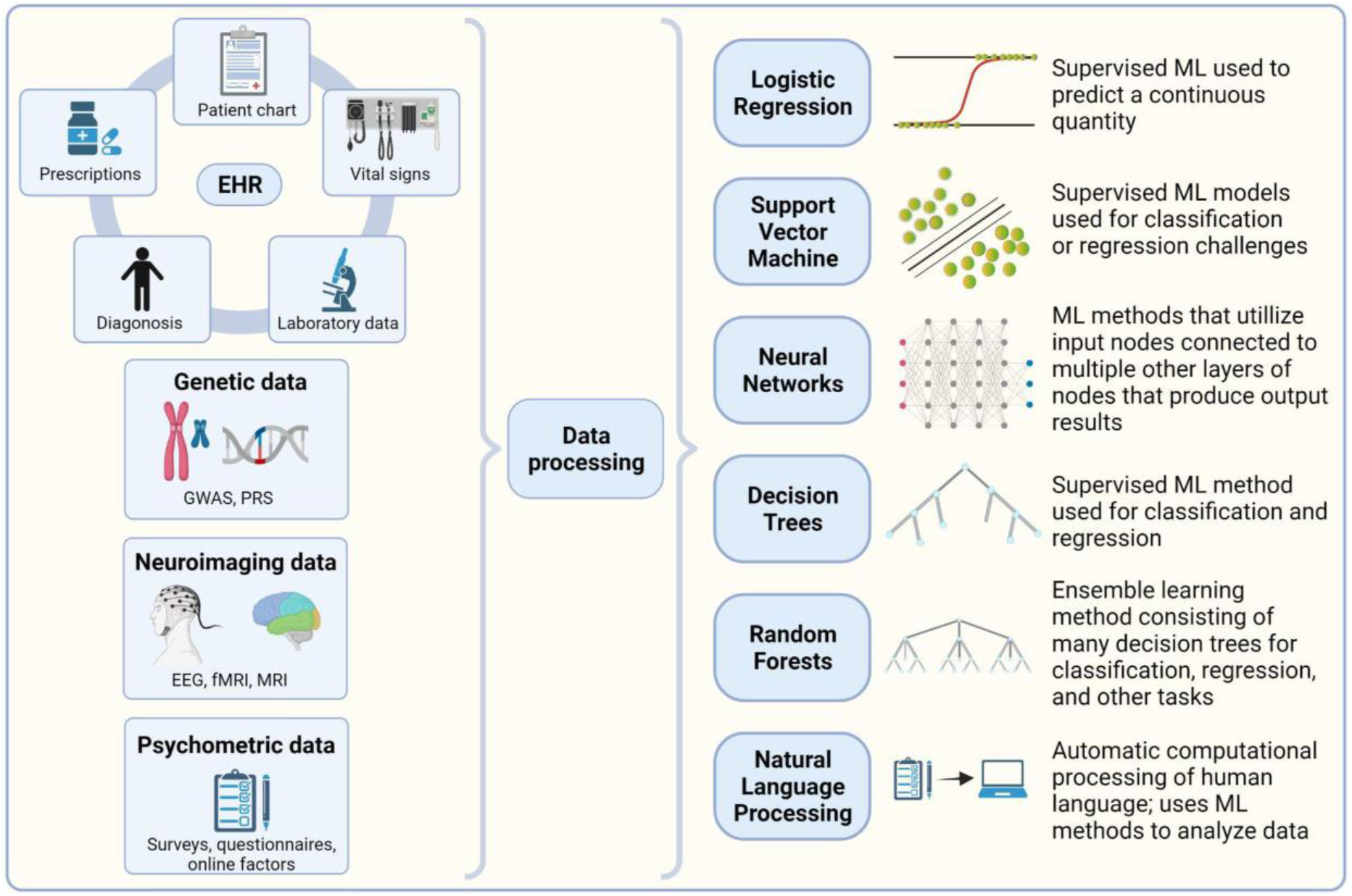
Overview of the types of data utilized and brief descriptions of the various applied machine learning methods in our selected studies for predictions on alcohol use disorder and related outcomes. Abbreviations: electroencephalography (EEG); electronic health record (EHR); functional magnetic resonance imaging (fMRI); genome wide association study (GWAS); machine learning (ML); magnetic resonance imaging (MRI); polygenic risk score (PRS).

### Screening for AUD

Five of the 70 studies included in this review aimed to simplify mass screening for AUD by using electronically available data for alcohol misuse identification. Among them, three studies used only data readily available in EHR, including structured data and non-structured clinical notes interpreted through natural language processing (NLP) for identifying alcohol misuse.(15–17) Highest performance was achieved using a logistic regression model (accuracy 0·91) with 25 input features collected via NLP.(17) The other two studies utilized social media platforms to screen for alcohol misuse-related behaviors. ML models were developed to classify binge drinking tweets(18) or Instagram posts(19) to identify risk of alcohol and/or drug misuse. However, largely due to ambiguities in assessing tweets, differentiating genuine and non-genuine users was difficult and affected the models’ accuracy (0·67 ± 0·05).

### Diagnosis/identification/discrimination of AUD

A substantial number of the studies focused on AUD diagnosis. These studies utilized a variety of data types, including laboratory measurements from blood tests, polysomnography, functional magnetic resonance imaging (fMRI), electroencephalography (EEG), and genotypic and/or phenotypic data. Single, indirect biomarker-based blood tests are not considered accurate for AUD diagnosis;(20) however, combinations of biomarkers used together with ML classified alcohol misuse successfully.(21–23) A combination of 2-5 biomarkers were analyzed using decision trees, artificial neural networks (NNs), and multivariate unequal dispersed class models. All three studies found mean corpuscular volume of red blood cells and gamma glutamyltransferase to be important features in their final models.

ML methods were developed to differentiate AUD and non-AUD individuals using neurobiological data. Polysomnographic data and neurobiological data were analyzed using feed-forward NNs, achieving 98% accuracy, sensitivity, and specificity, and demonstrating that alcohol addiction correlates with deep sleep impairment.(24) This research also reported that importance of features varied between men, women, and mixed sex groups.(24)

Most ML-based studies in the field of neuroimaging for AUD diagnosis fell into two major categories: identifying AUD from differences in structural data, network connectivity, or brain volume based on neuroimaging(25–32), and identifying AUD from EEG data where features were derived from electrical signals.(33–44) ML models based on neuroimaging was predominantly trained with MRI data collected from dozens of participants with or without AUD or known alcohol-dependent behaviors.(25–32) Of these studies, several have developed and tested models based solely on structural and/or network connectivity data collected by fMRI, thereby identifying potential neuroimaging markers of alcohol dependency detected by standard imaging techniques.(25,28,30) Others have supplemented neuroimaging with other data sources and variables to refine diagnostic accuracy; these included neuropsychological scoring (variables of memory span and scored results of the visual span test), psychosocial factors related to behavior and environment (history of substance use, relationship and friendship status, personality traits, and emotional traits), and human immunodeficiency virus-AUD comorbid status.(26,27,32) Two neuroimaging studies by Wang et al. outperformed other contemporary studies with similar dataset sizes; one study detected AUD by identifying features weighted by a convolutional NN trained from 160 images and tested on 159,(29) another study assigned feature weights via a transfer learning model trained on 100 images and tested on 135.(31)

AUD diagnostic models that are based on EEG data appear to outperform those based on neuroimaging data, although a direct comparison is not possible because of variation in reported performance metrics and incomplete records of data missingness. Utilizing EEG data, relatively high diagnostic accuracy ranging from 80-99% was reported, in comparison to neuroimaging studies ranging from 67-87% accuracy, with the exception of Wang et al.’s imaging-based algorithms achieving 97% accuracy.(29,31,33–46) Studies using EEG features have consistently shown strong results when employing support vector machine (SVM) or least-squares support vector machine (LS-SVM) architecture, although methods for extracting and decomposing EEG features have differed widely.(33,34,37,38,40–46) Other studies developed and tested multiple models from the same datasets of extracted EEG features, with SVM, LS-SVM, and convolutional NN-based architectures generally yielding the most reliable results.(37,39,42,44–46) Strong performance was also demonstrated from 10-fold cross-validation test data collected from single channels and even single electrodes, with Hussain et al. reporting AUROC values of 0·976 to 0·998 from multi-scale entropy- and fast multi-scale entropy-based models using data from the C3 central electrode and Kumar et al. reporting an SVM-based model’s classification accuracy of 88% using data from only the F4 channel.(37,45)

ML using genotypic and phenotypic data has been useful to comprehensively analyze large datasets in AUD research. Phenotypic analysis was explored by Li et al. and Falk et al. using NNs for AUD diagnosis, where features of drinking patterns and its effects and phenotypic variables resulted in 95% prediction accuracy.(47,48) Yu et al. studied microsatellite markers to evaluate the linkage between AUD and specific genomic regions.(49) Chen et al. used single nucleotide polymorphisms (SNPs) combined with age, education level, and marital status to create a personalized approach to discriminate between alcohol dependent and non-alcohol dependent patients.(50) Another study demonstrated that salivary microRNAs (miRNAs) may potentially be used to recognize alcohol dependence (AD).(51)

### Predicting AUD severity

Beyond AUD diagnosis, assessment of AUD severity is a critical area of research. Fede et al. examined the association of AUD severity as measured via Alcohol Use Disorders Identification Test (AUDIT) and differences in patient neurobiology by applying ML to MRI scans.(52) Their results indicated an ML model based on resting state-connectivity features can best distinguish varying levels of AUD severity, and can potentially be used as neuroimaging biomarkers for clinical evaluation.

### Risk factors for alcohol use

Analysis of risk factors for AUD using ML was also investigated. Particularly, in genetics, random forest analyses with and without X chromosome data produced variable importance estimates for X chromosome variants when biological sex was associated with AUD.(53) Sex differences in the heritability of alcohol misuse was previously demonstrated.(54,55) However, most ML models incorporating genetic data did not correctly model the effects of the X chromosome SNPs for several reasons: X chromosome data were routinely excluded from GWAS(56); the number of X chromosome copies is confounded with sex necessitating special analysis; and incorporation of X chromosome inactivation into statistical analysis remains difficult because its mechanisms are not yet fully understood.(57)

### Predicting future alcohol use

Predicting future development of AUD using ML is an active area of research. A comprehensive prospective study conducted by King, et al., developed a logistic regression model to predict future occurrence of hazardous drinking.(58) The model identified sex, age, country, AUDIT score, panic syndrome, and lifetime alcohol problem as risk factors, and resulted in a c-index value of 0.78 in the external validation test set of non-AUD Chilean drinkers. In a separate study, SNPs were used to predict who is likely to develop AUD and identify biomarkers that indicate a predisposition to AUD.(59) Results demonstrated that models that combined genetic and electrophysiology features achieved higher accuracy compared to one-dimensional models.

### Prediction of treatment outcomes

While majority of the studies included in this review aimed to predict and diagnose alcohol misuse, 19 studies focused on prediction of AUD treatment outcome,(60–78) and one study was directed at identifying treatment seeking AUD patients.(79) Several studies developed ML methods to predict relapse,(60,61,64–66,69) most of which reported demographic data, behavioral and psychological measures, alcohol consumption, and dependence severity measures as important predictors. Methods to predict AUD treatment outcomes after cognitive behavioral therapy reported moderate accuracy.(60,61,69) Symons et al. 2019(61) and 2020(60) reported that their ML models outperformed clinical judgment for predicting treatment outcome when given the same data, including drinking-related measures, demographic, and psychological assessment data. However, these models yielded poor sensitivities, ranging from 8 - 43%. Treatment outcome predictions based on data from a self-help intervention internet diary(62) or a smartphone application(68) have demonstrated high accessibility.

ML methods were used to make accurate predictions of health-related outcomes for AUD patients as well, including risk of withdrawal seizures and mortality. Models assessing risk of alcohol withdrawal seizures using homocysteine data(70,71) and prediction of alcohol withdrawal severity(72) may help clinicians determine treatment course. Survival prediction models using ML algorithms were also developed for alcohol-dependent patients with severe liver disease(73) or drug intoxication(74) upon hospital admission based on demographic data, clinical variables, and medical history. The ML model developed by Lapuerta et al.(73) outperformed the Maddrey score(80) in predicting survival of severe liver disease patients (ROC area of 81·5% vs. 73·8%).

ML approaches have also impacted the prediction of subgroups of individuals who may benefit from specific pharmacotherapy.(75–78) Laska et al. focused on identifying responders to Gabapentin Enacarbil Extended-Release using demographic, substance use indicators, and psychiatric characteristics.(77) Other researchers utilized genetic information: Hou et al. and Wei et al. examined SNPs to identify subgroups that will benefit from ondansetron treatment for AUD.(75,76) Lin et al. investigated how *OPRM1* promoter CpG site methylation affects relapse in individuals taking naltrexone for alcohol dependence.(78)

### AUD in adolescents

Consumption of alcohol during adolescence may disrupt neurodevelopmental trajectories.(81) Several ML studies have incorporated multifactorial neurological characteristics as well as demographic, behavioral, cognitive, and clinical features to identify risk factors and predict AUD in adolescents. Squeglia et al. employed ML on multiple data sources in a longitudinal study of substance-naive adolescents to identify predictors of alcohol use by age 18, half of which were sMRI and fMRI variables.(82)

Additional studies used multiple algorithms to investigate the interaction of features that are specific for alcohol use in adolescents. These studies identified risk factors (83–85) or predicted alcohol misuse (86) in adolescents using combinations of demographic, psychopathological and personality data, socioeconomic data, and cognitive measures. The influence of peers and parents, and respondents’ sex (male) had the highest impact on substance use initiation during childhood for Mexican youths.(84) In contrast, personality and psychopathology factors yielded the highest prediction accuracy indices in a prospective study of Canadian and Australian adolescents.(86) Particulary, personality traits such as disorderliness and extravagance was correlated with both current and future adolescent binge drinkers.(85)

Neuroimaging was also used in a unique study that investigated potential biomarkers of resilience to alcohol misuse in youths. Weidacker et al. concluded that grey matter myelinations (myeloarchitecture) could be a potential protective biomarker, as greater baseline myeloarchitecture predicted a lower risk for harmful alcohol use at two-year follow-up.(87)

Impulsivity is another aspect of interest for AUD prediction in adolescents. Sex-dependent differential trajectories of impulsivity in high school students may increase an individual’s susceptibility to substance use disorder, including alcohol.(88) Ruan et al. found that impulsivity did not decline [as in normal development] for adolescents who initiated binge drinking.(89) Other studies showed that AUD could be predicted using impulsivity variables based on brain activity, personality, and psychological factors(90) and that not all facets of impulsivity may be associated with AUD.(91)

Overall, 66 of the 70 the studies included in this review exhibited a moderate or high risk of bias, predominantly from a lack of external validation in algorithm development, and missing data (Supplementary Table 4). 25 (35·7%) of studies included in this review were scored as a high risk of bias, 41 (58·6%) as a moderate risk of bias, and 4 (5·7%) as a low risk of bias in our analysis.

## DISCUSSION

This systematic literature review presents the current state of research for ML for alcohol misuse, AUD, and AUD-associated consequences. To our knowledge this is the first systematic review to provide an overview of ML-based techniques for alcohol misuse and AUD. While it is not possible to directly compare all of the included studies because of differences in the parameters, cohorts, type of data, and assessment approaches used, overall this overview demonstrates encouraging results for the use of ML for the identification and treatment of individuals with AUD-related predictions. This review highlights research that used readily available EHR data and neuroimaging data to accurately identify and diagnose individuals with AUD, including adolescents at high risk. Several genetic-based studies demonstrate the potential of using ML to analyze genetic variations for personalized treatments or prediction of the risk of AUD. We also identified several ML-based studies that use clinical measures to predict effectiveness of a particular treatment plan, risk stratify patients to improve health outcomes, and determine potential relapse. These diverse ML-based studies identified numerous significant variables for AUD and AUD-related predictions (Figure 3).

**Figure 3.**
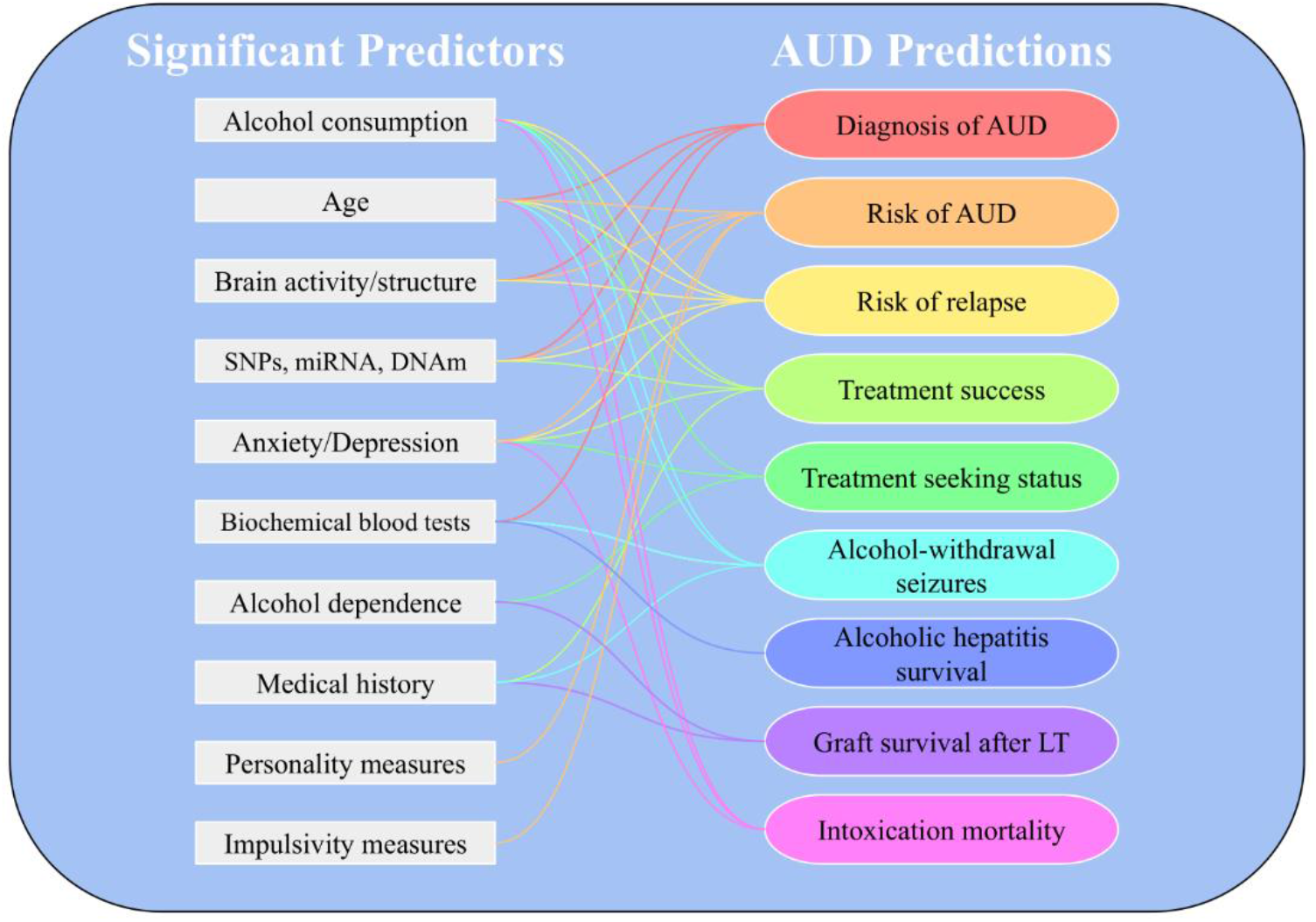
Summary of significant predictors for alcohol use disorder-related predictions identified in our systematic review. Abbreviations: Alcohol use disorder (AUD); DNA methylation (DNAm); liver transplantation (LT); microRNA (miRNA).

The use of ML to assist in diagnosis and risk assessment in healthcare is potentially powerful, given the rich and variable data sources available. Use of multiple data sources in ML, including neuroimaging, electrophysiological, cognitive, genetic, socioecological, psychological, and demographic data, facilitates better classification accuracy in numerous studies in comparison to single dimensional data.(85,92) Multi-dimensional modeling may better identify the underlying pathophysiology of a complex disease such as AUD, and may lead to improved prediction methods for diagnosis, risk stratification of patients, and healthcare resource allocation. ML may reveal data relationships or potential predictors not previously known in the field, such as specific measures of impulsivity, SNPs, or laboratory measures associated with AUD. These findings can be used to identify significant areas to investigate for future AUD research to address current barriers, such as treatment utilization rates and AUD relapse.

Implementing ML methods may reach larger, currently underserved patient populations. ML-based tools that use readily available EHR data may cost-effectively diagnose AUD with no additional burden on the clinician and may increase the low AUD screening rates.(6) ML in combination with other modern technologies like social media and internet based self-help tools may be useful to those who might otherwise go undiagnosed or not seek treatment. Top reported reasons for not seeking AUD treatment are a ‘lack of problem awareness’, ‘stigma or shame’, ‘encounter barriers’, and ‘cope alone’.(93) Convenient ML-based tools that rely on easily accessible EHR data or internet data such as the ones described in this review have the potential to overcome these limitations by screening more of the public or providing a more anonymous means of receiving care for those whose shame prevents them from seeking treatment.

Genetic and neuroimaging-based studies showed considerable innovation in the field because they may provide clinicians with methods to predict AUD and treatment outcomes without relying on largely biased and unreliable self-reported patient behavior.(94) As genetic and neuroimaging techniques develop and data become more widely available,(95) the accuracy of predicting AUD with increasingly innovative ML approaches will likely continue to improve over time. These methods can be efficiently integrated into contemporary healthcare delivery information systems. The outstanding sensitivity and high to very high specificity of the neuroimaging-based algorithms (MRI-based methods sensitivity: 97%, specificity: 97%(29,31); EEG-based methods sensitivity: 90-99·99%, specificity: 82-99·97%(33–35,38,40,41,44)) show their potential to assist clinicians in diagnosing and intervening in AUD development.

These findings also emphasize that ML methods are well equipped to address the multi-faceted nature of alcohol use and the complexity of pathological behavior in adolescents. Diverse risk factors are potentially influential in children and adolescents at different ages, where they exhibit nuanced individual differences in maturity and development. As the use of ML techniques to develop computerized tests is an emerging trend in mental health,(96) these studies affirm the potential of using ML algorithms in the development of versatile computerized tests for the broad screening of high-risk adolescents in both clinical and school settings. From a clinical perspective, this would allow the early identification and intervention of at-risk adolescents, conceivably mitigating future harm caused by alcohol misuse.

Most of our selected studies exhibited a high or moderate risk of bias. Future studies in this field may aim to reduce bias by assessing algorithm performance in external dataset validations. Some algorithms did not achieve sufficient performance to be implemented in healthcare. Future work may also include refining ML techniques and enhanced research efforts in this field to work towards integration of these models into routine clinical care.

There are several limitations in our systematic review. The screening process included determining whether a study’s methods were considered to be ML or conventional statistics at our discretion, potentially introducing bias in screening. The risk of bias assessment and potential bias sources were also manually determined for each study. Several of the included studies did not use a comparator for reference on ML models’ performance, therefore it was not possible to determine if those studies improved predictions compared to current methods.

## CONCLUSIONS

Here we presented a systematic literature review on the use of ML-based techniques for AUD and AUD-related consequences concerning health, treatment, recovery, and prevention. This systematic review summarizes the current state of research and identifies future research opportunities for AUD. The research presented has established the versatility and usefulness of ML in AUD, utilizing data from highly diverse sources (including EHR, neuroimaging, genetic, and psychometric data) to generate accurate classification and predictions. Although further investigation and refinement is needed, available evidence suggests that ML-based tools for clinical practice in AUD show promise.

## Data Availability

Data are based on the reported findings of published articles listed in the tables and are available online.

## AUTHOR CONTRIBUTIONS

MH performed full text screening, created figures, extracted data, and contributed to the writing. AS conceived the study, designed the search strategy, performed title/abstract screening, performed full text screening, and edited the manuscript. MMA performed full text screening, created tables and figures, extracted data, and contributed to the writing. ZI performed title/abstract screening, extracted data, and contributed to the writing. DE extracted data and contributed to the writing. JH provided oversight and leadership responsibility. All authors verify that they had full access to the data in this systematic review.

## ACKNOWLEDGMENTS

The authors would like to gratefully acknowledge Dr. Nicole S. Zelin and Dr. Abigail Green-Saxena for their assistance with title and abstract screening. Figure 2 was created with BioRender.com.

