## Supplementary Table for "Machine Learning Applications and Advancements in Alcohol Use Disorder: A Systematic Review"

### Supplementary Materials

**Supplementary Table 1.** Search strategy used for identification of articles on Embase and PubMed Central.

| Database | Search Strategy |
|---|---|
| Embase | ('alcoholism'/exp OR 'alcohol consumption'/exp OR 'alcohol use disorders identification test'/exp OR 'alcohol psychosis'/exp OR 'alcohol liver disease'/exp OR 'alcohol withdrawal syndrome'/exp OR 'alcoholic cardiomyopathy'/exp OR 'alcohol abuse'/exp OR 'alcoholic pancreatitis'/exp OR 'fetal alcohol syndrome'/exp OR 'alcohol intoxication'/exp OR 'alcohol abstinence'/exp OR 'acamprosate' OR 'disulfiram' OR 'naltrexone' OR 'varenicline' OR ‘topiramate’ OR ‘gabapentin’) AND ('machine learning'/exp OR 'artificial intelligence'/exp OR 'decision support system'/exp) |
| PubMed Central | (“alcohol related disorders”[Mesh] OR “alcoholics”[Mesh] OR “alcohol withdrawal delirium”[Mesh] OR “alcohol withdrawal seizures”[Mesh] OR “drinking behavior”[Mesh] OR “alcohol deterrents”[Mesh]) AND (“algorithms”[Mesh] OR “machine learning”[Mesh] OR “natural language processing”[Mesh] OR “neural networks, computer”[Mesh] OR “decision support systems, clinical”[Mesh]) |

**Supplementary Table 2.** Overall scoring for risk of bias assessment.

| Risk of Bias Score | Rationale |
|---|---|
| "LOW" | Data source: multiple sources, or a national database. Missing data: none. External validation: performed. |
| "MODERATE" | Missing data: moderate amount of data missing. All categories: not specified. |
| "HIGH" | Data source: data collection from only one clinical site. Missing data: high amount of data missing. External validation: no external validation performed. |
| Overall Risk of Bias Score: "LOW" | No categories with "HIGH" risk of bias. |
| Overall Risk of Bias Score: "MODERATE" | One category with "HIGH" risk of bias, or one or two categories with "MODERATE" risk of bias. |
| Overall Risk of Bias Score: "HIGH" | Two or more categories with "HIGH" risk of bias, or three or more categories with "MODERATE" risk of bias. |

**Supplementary Table 3.** Study design details of selected articles.

| First Author, Year | Type of ML algo | Sample Size | Data Type | Ethnicity/race | Age | Gender | Comparator or competitor |
|---|---|---|---|---|---|---|---|
| Screening | | | | | | | |
| Afshar, 2019 | NLP; LR, PAC, SVM, SGD | 1422 | EHR, including clinical notes | 54% White; 18% Hispanic | Range ≥18 | 71% male | Rules based pre-selected keyword approach (keywords chosen by content experts); AUDIT |
| To, 2020 | LR with LASSO; NLP | 2422 | EHR, including clinical notes | 58% White | Range ≥18 | N/S | None |
| Bonnell, 2020 | LR, SVM; NN, k-nearest neighbors, DT, RF (RF used for final model) | 43545 | Demographic, clinical and laboratory information | N/S | Range ≥18 | 49% male | AUDIT-C |
| Hassanpour, 2019 | CNN and LSTM | 2287 | Instagram pictures and captions | White, Black, Asian, Hispanic | Range 18+ | Males and females | None |
| Crocamo, 2020 | SVM and RF | 180 | Tweet content | N/S | N/S | N/S | None |
| Diagnosis/identification/discrimination of AUD | | | | | | | |
| Maurelli, 1998 | Genetic algorithm BEAGLE; 7 ANNs; Metasystem taking outputs from ANNs to produce new results | 113 | Blood markers: MCV, ALKPHOS, ALT, AST, GGT | N/S | N/S | N/S | None |
| Dinevski, 2011 | DT | 482 | Blood markers: MCV, GGT, GLDH, AST, ALT | N/S | Range 18-65 | 82-84% male | Individual biomarkers |
| Pirro, 2013 | UNEQ model | 423 | Blood markers: AST, ALT, GGT, MCV, CDT | N/S | Range 19-74 | ~80% male | Individual biomarkers |
| Lewenstein, 2020 | ANN | 172 | Polysomnography (EEG, EOG, EMG) | N/S | Range 30-46 | 50% male | None |
| Guggenmos, 2018 | Weighted robust distance-to-centroid classifier | 216 | MRI | N/S | Range 20-65 | 84% male | None |
| Kamarajan, 2020 | RF | 60 | fMRI | 42% Black; 15% White; 35% Asian; 2% American Indian | Mean 41 (AUD); 27 (control) | 100% male | None |
| Gowin, 2020 | RF | 486 | fMRI and self-reported psychological / substance use history | AUD: 83% White; 9% Black; 7% Hispanic; Control: 69% White; 20% Black; 9% Hispanic | Mean 28±3 (AUD); 29±4 (control) | 72% (binge-drinkers); 33% (non-bingers) | None |
| Hahn, 2020 | LR and SVM | 2034 | MRI | N/S | Mean 45 (test-AUD); 42 | 57% male | None |

| | | | | | | | |
|---|---|---|---|---|---|---|---|
| | | | | | (test-control) | | |
| Wang, 2019 | AlexNet transfer learning model | 379 | MRI | N/S | N/S | N/S | None |
| Zhu, 2018 | RF | 92 | fMRI | AUD: 54% White; 41% Black; 4% Asian; Control: 63% White; 28% Black; 9% Asian | Mean 40±10 (AUD); 32 ±9 (control) | 72% male (AUD), 57% male (control) | None |
| Wang, 2017 | CNN | 235 | fMRI | N/S | N/S | N/S | None |
| Adeli, 2019 | LR | 549 | fMRI | 7% Asian; 21% Black; 61% White; 11% Other | Mean 48 ±10 (AUD); 47±14 (control); 51±9 (HIV); 52±7 (AUD+HIV) | 54% male (control), 66% male (AUD), 69% male (HIV), 63% male (AUD+HIV) | None |
| Bae, 2017 | SVM | 60 | EEG | N/S | N/S | N/S | None |
| Acharya, 2012 | SVM | 122 | EEG | N/S | N/S | N/S | None |
| Mehla, 2020 | SVM | 122 | EEG | N/S | N/S | N/S | None |
| Hussain, 2018 | k-D tree | 58 | EEG | N/S | Mean 36±5 (AUD); 26±3 (control) | N/S | None |
| Padma Shri, 2017 | k-NN | 1200 | EEG | N/S | N/S | N/S | None |
| Mumtaz, 2017 | LR | 45 | EEG | N/S | Mean 55±13 (AUD); 43±16 (control) | N/S | None |
| Mumtaz, 2018 | SVM | 60 | EEG | N/S | N/S | N/S | None |
| Kumar, 2016 | SVM and FCM | 40 | EEG | N/S | Range 32-38 | 100% male | None |
| Kumar, 2015 | SVM and FCM | 40 | EEG | N/S | Mean 35 | 100% male | None |
| Zhang, 2020 | CNN and SVM | 125 | EEG | N/S | Mean 36±5 (AUD); N/A (controls) | 100% male | None |
| Anuragi, 2019 | LS-SVM | 122 | EEG | N/S | N/S | N/S | None |
| Anuragi, 2020 | LS-SVM | 122 | EEG | N/S | N/S | N/S | None |
| Faust, 2013 | FSC | 122 | EEG | N/S | N/S | N/S | None |
| Zhu, 2014 | HVGE | 122 | EEG | N/S | N/S | N/S | None |
| Chen, 2020 | GS-SVM, CNN, CNN-LSTM | 297 | SNPs, demographic information | Chinese | Range Adults | 100% male | None |
| Yu, 2005 | SVM | 1204 | Microsatellite markers | N/S | N/S | N/S | None |
| Rosato, 2019 | RF | 120 | microRNA | 53% European American; 47% | Range 18+ | 47% male | None |

| | | | | | | | |
|---|---|---|---|---|---|---|---|
| | | | | African American | | | |
| Falk, 2005 | ANN | 650 | Risk factors, drinking habits and phenotypic measurements | N/S | N/S | N/S | None |
| Li, 1999 | ANN | 699 | Phenotypic ERP300 data; genotypic data; biomarker: MAO; gender | N/S | N/S | N/S | None |
| Predicting AUD severity | | | | | | | |
| Fede, 2019 | RF | 59 | fMRI | N/S | Range 22-60 | 68% male (primary); 50% male (validation) | None |
| Risk factors for alcohol use | | | | | | | |
| Winham, 2016 | RF | 3829 | SNPs | 69% European American; 31% African American | Range 18-77 | 46% male | None |
| Predicting future alcohol use | | | | | | | |
| King, 2011 | LR | 8655 | Demographic, medical and psychological history, living conditions, AUDIT questionnaire; behavioral questions; alcohol, drugs and cigarette use; history of abuse; religious beliefs; family history of mental health; stress, anxiety and panic syndrome; List of Threatening Life Experiences Questionnaire; experiences in discrimnation; social support | European set: 97% White European Chilean set: 100% Non-white | Range 18-75 | European set: 31% male; Chilean set: 24% male | None |
| Kinreich, 2019 | SVM | 656 | EEG, SNPs | European American and African American | Range 12–30 | 57% male | None |
| Prediction of Treatment Outcomes | | | | | | | |
| Symons, 2020 | RF, BayesNet, NB, SPegasos, RBFN, SMO, REPTree, DTNB, FURIA, Decision table | 2236 | Demographic, medical history, alcohol dependence measures, and psychological measures | N/S | Range 18–76 | 65-71% male | 10 psychologists and traditional linear regression |
| Symons, 2019 | FURIA, ADT, LWL, JRip, RIDOR, SMO, REPTree, SPegasos, Decision Table, RBFNM, BayesNet, DTNB, BFTree | 830 | Demographic and psychometric assessment data, medical history and drinking measures | N/S | Range 18-76 | N/S | 10 addiction therapists |

| | | | | | | | |
|---|---|---|---|---|---|---|---|
| Lindner, 2020 | RF | 607 | Drinking measures | N/S | N/S | Males and females | None |
| Sekutowicz, 2019 | linear SVM | 188 | fMRI | N/S | Mean 45 | 84% male | None |
| Seo, 2015 | NB, SVM, LVQ | 46 | sMRI and fMRI | N/S | Average 40 | 65% male | ML models that used clinical data including demographics, medical history, and socioeconomic status. |
| Winterer, 1998 A | ANN | 78 | Quantitative EEG | N/S | Mean 45-48 | 58% male | None |
| Winterer, 1998 B | ANN | 78 | Quantitative EEG | N/S | Mean 45-48 | 58% male | ML models that used clinical data including demographic data, laboratory, psychiatric and neurological measurements |
| Satapathy, 2020 | LR | 241 | Demographic, medical and previous rehabilitation history | 73% Caucasian; 17% Black; 8% Hispanic; 0.4% Other | Mean 53 | 81% male | None |
| Chih, 2014 | Bayesian network model | 152 | Behavioral and psychological measures, substance use, and lapse history | 83% Caucasians | Range 20-64 | 62% male | None |
| Connor, 2007 | Bayesian network model and DTs | 139 | Demographic, medical history, dependence severity, and drinking-related and psychological measures. | N/S | N/S | Males and females | Discriminant analysis |
| Kurth, 2001 | LR and ANN | 49 | Demographic, laboratory values, and alcohol withdrawal assessment data | N/S | Range 27-66 | 86% male | None |
| Hillemacher, 2012 | RF | 200 | Demographic, medical history, and laboratory variables | N/S | Mean 45-48 | 81% male | None |
| Burkhardt, 2020 | SVM | 1144 | Laboratory variables, sociodemographic measures, and medical history | N/S | Mean 43 | 73-79% male | None |
| Lapuerta, 1997 | ANN and LR | 144 | Laboratory values and clinical variables | 66% Hispanic; 25% White; 5% African-American | Mean 40 | 83% male | Maddrey score |
| Choi, 2020 | LR, DT, and MLP | 8937 | Demographic, psychological and behavioral data | East Asian | <65; 65+ | Males and females | None |
| Hou, 2015 | IT, TRM, and VT | 251 | SNPs | Hispanic and others | Range 20-78 | N/S | None |
| Wei, 2020 | IT-LT | 251 | SNPs | Hispanic and others | Range 20-78 | N/S | None |
| Laska, 2020 | RF | 338 | Demographic, substance use indicators, and psychiatric characteristics | 65-77% White; 16-23% Black | Mean 50 | 66% male | None |

| | | | | | | | |
|---|---|---|---|---|---|---|---|
| Lin, 2020 | RF | 93 | Demographic, OPRM1 promoter region DNA methylation | 44% African American; 36% European American | Range 50-51 | 100% male | None |
| Predicting treatment seeking behavior | | | | | | | |
| Lee, 2019 | ADT, RF, RT | 1114 | Behavioral and psychological data | 39-73% African American; 20-52% Caucasian; 7-13% Other | Range 21-69 | 64-77% male | None |
| AUD in Adolescents | | | | | | | |
| Squeglia, 2017 | RF | 137 | sMRI, fMRI, demographic, neuropsychological data | 68% Caucasian; 32% N/S | Range 12-16 | 56% male | None |
| Whelan, 2014 | Voxelwise LR, Regularized LR | 2000 | sMRI, fMRI, demographic, neuropsychological data | Caucasian | Range 14-18 | 48% male | None |
| Afzali, 2019 | LR, SVM, RF, NN, Lasso Reg, Ridge Reg, Elastic-net | 6016 | Demographic, psychopathological, personality, risk behaviors, cognitive | N/S | Range 12-16 | N/S | None |
| Ruan, 2019 | SVM | 2000 | rsMRI, fMRI, rsFC, SNPs, personality, cognitive | Caucasian | Range 14-19 | 49% male | None |
| Vázquez, 2020 | Elastic net, k-nearest neighbors, NN, RF | 191880 | Demographic, socioecological, personality, | Mexican | Range 10-12 | 51% male | None |
| García, 2009 | ANN, Decision trees, Naive bayes | 7030 | Personality variables from survey | Mexican | Range 14-18 | N/S | None |
| Weidacker, 2020 | RVR | 3064 | sMRI, fMRI, demographic, behavioural data | N/S | Range 15-26 | 67% male | None |
| O'Halloran, 2020 | Elastic net | 79 | EEG, demographic, personality, drinking behavior, task-based tests | N/S | Range 18+ | 49% male | None |
| O'Halloran, 2018 | Elastic net | 106 | Demographic, behavioural, personality | N/S | Range 18-21 | 66% male | None |
| Martinez-Loredo, 2018 | Binary logistic regression | 1239 | Demographic, behavioural, personality | Spaniards | Range <15 | 54% male | None |

Abbreviations: Alanine aminotransferase (ALT); Alcohol use disorder (AUD); Alcohol Use Disorder Identification test (AUDIT); Alkaline phosphates (ALKPHOS); Alternating decision tree (ADT); Artificial neural network (ANN); Aspartate aminotransferase (AST); Biologic Evolutionary Algorithm Generating Logical Expressions (BEAGLE); Best-first decision tree (BFTree); Carbohydrate-deficient transferrin (CDT); Convolutional neural network (CNN); Decision table naive bayes (DTNB); Decision tree (DT); Electroencephalographic (EEG); Electromyogram (EMG); Electronic health record (EHR); Electrooculograms (EOG); Event-related potential (ERP); Functional MRI (fMRI); Fuzzy C-mean (FCM); Fuzzy sugeno classifier (FSC); Fuzzy unordered rule induction algorithm (FURIA); Gamma-glutamil transpeptidasis (GGT); Glutamate dehydrogenase (GLDH); Horizontal visibility graph entropy (HVGE); Interaction tree (IT); Interaction tree for longitudinal trajectories (IT-LT); Learning vector quantization (LVQ); Least Absolute Shrinkage and Selection Operator (LASSO); Locally weighted learning (LWL); Logistic regression (LR); Long short-term memory (LSTM); Magnetic resonance imaging (MRI); Mean corpuscular volume (MCV); Monoamine oxidase (MAO); Multilayer perceptron model (MLP); Multivariate unequal dispersed classes (UNEQ); Naïve Bayes (NB); Natural language processing (NLP); Neural network (NN); Passive Aggressive Classifier (PAC); Radial basis function neural networks (RBFN); Random forest (RF); Random tree (RT); Reduced-error pruning tree (REPTree); Relevant Vector Regression (RVR); Resting-state functional connectivity (rsFC); Ripple down rule learner (RIDOR); Sequential minimal optimization (SMO); Single nucleotide polymorphism (SNP); Support vector machine (SVM); Traditional regression method (TRM); Virtual twins (VT)

**Supplementary Table 4.** Risk of bias assessment of selected studies

| First Author, Year | Sample source | Missing data | External Validation | Other sources | Overall risk |
|---|---|---|---|---|---|
| Screening | | | | | |
| Afshar, 2019 | HIGH; 1 source | LOW | HIGH: no | MODERATE: based on clinical notes analyzed from 1 center, and language variation in clinical notes in other locations may vary | HIGH |
| To, 2020 | HIGH; 1 source | LOW | HIGH: no | MODERATE: based on clinical notes analyzed from 1 center, and language variation in clinical notes in other locations may vary | HIGH |
| Bonnell, 2020 | LOW; Large dataset | LOW | HIGH: no | HIGH: did not use EHR to build algorithm, but target is to use tool on EHR, which has a much higher %age of missing data | HIGH |
| Hassanpour, 2019 | HIGH; Social media platform | MODERATE: N/S | HIGH: no | LOW: N/S | HIGH |
| Crocamo, 2020 | MODERATE; Social media platform | MODERATE: N/S | HIGH: no | HIGH: ambiguities in assessing tweets | HIGH |
| Diagnosis/identification/discrimination of AUD | | | | | |
| Maurelli, 1998 | HIGH; 1 source | MODERATE: N/S | HIGH: no | LOW: N/S | HIGH |
| Dinevski, 2011 | HIGH; 1 site | LOW | HIGH: no | MODERATE: Control patients and patients with alcohol dependence syndrome from different sources | HIGH |
| Pirro, 2013 | MODERATE; 2 sites | LOW | HIGH: no | LOW: N/S | MODERATE |
| Lewenstein, 2020 | HIGH; 1 source | MODERATE: N/S | HIGH: no | LOW: N/S | HIGH |
| Guggenmos, 2018 | LOW; Multisite study | MODERATE: N/S | LOW: yes | LOW: N/S | MODERATE |
| Kamarajan, 2020 | MODERATE; 1 source but from larger study | MODERATE: N/S | HIGH: no | LOW: N/S | MODERATE |
| Gowin, 2020 | MODERATE; 1 source but sampled from larger study | MODERATE: N/S | HIGH: no | LOW: N/S | MODERATE |

| | | | | | |
|---|---|---|---|---|---|
| Hahn, 2020 | LOW; Multisite study | MODERATE: N/S | LOW: yes | LOW: N/S | LOW |
| Wang, 2019 | HIGH; 1 source | MODERATE: N/S | LOW: yes | LOW: N/S | MODERATE |
| Zhu, 2018 | LOW; NIH recruited | MODERATE: N/S | HIGH: no | LOW: N/S | MODERATE |
| Wang, 2017 | LOW; 3 sites | MODERATE: N/S | HIGH: no | LOW: N/S | MODERATE |
| Adeli, 2019 | | MODERATE: N/S | HIGH: no | LOW: N/S | MODERATE |
| Bae, 2017 | LOW; Repository for large datasets | MODERATE: N/S | HIGH: no | LOW: N/S | MODERATE |
| Acharya, 2012 | LOW; Repository for large datasets | MODERATE | HIGH: no | LOW: N/S | MODERATE |
| Mehla, 2020 | LOW; Repository for large datasets | MODERATE | HIGH: no | LOW: N/S | MODERATE |
| Hussain, 2018 | LOW; Repository for large datasets | MODERATE: N/S | HIGH: no | LOW: N/S | MODERATE |
| Padma Shri, 2017 | HIGH; 1 site | MODERATE: N/S | HIGH: no | LOW: N/S | HIGH |
| Mumtaz, 2017 | HIGH; 1 site | MODERATE: N/S | HIGH: no | LOW: N/S | HIGH |
| Mumtaz, 2018 | MODERATE; N/S | MODERATE: N/S | HIGH: no | LOW: N/S | HIGH |
| Kumar, 2016 | HIGH; 1 source | MODERATE: N/S | HIGH: no | LOW: N/S | HIGH |
| Kumar, 2015 | HIGH; 1 source | MODERATE: N/S | HIGH: no | LOW: N/S | HIGH |
| Zhang, 2020 | LOW; Repository for large datasets | MODERATE: N/S | HIGH: no | LOW: N/S | MODERATE |
| Anuragi, 2019 | LOW; Repository for large datasets | MODERATE | HIGH: no | LOW: N/S | MODERATE |
| Anuragi, 2020 | LOW; Repository for large datasets | MODERATE | HIGH: no | LOW: N/S | MODERATE |
| Faust, 2013 | LOW; Repository for large datasets | MODERATE | HIGH: no | LOW: N/S | MODERATE |
| Zhu, 2014 | LOW; Repository for large datasets | MODERATE | HIGH: no | LOW: N/S | MODERATE |
| Chen, 2020 | HIGH; 1 site | MODERATE: N/S | HIGH: no | LOW: N/S | HIGH |
| Yu, 2005 | LOW; 9 sites across the US | MODERATE: N/S | HIGH: no | LOW: N/S | MODERATE |
| Rosato, 2019 | HIGH; 1 site | MODERATE: N/S | HIGH: no | LOW: N/S | HIGH |
| Falk, 2005 | LOW; 9 sites across the US | MODERATE: N/S | HIGH: no | LOW: N/S | MODERATE |
| Li, 1999 | LOW; 9 sites across the US | MODERATE: N/S | HIGH: no | LOW: N/S | MODERATE |
| Predicting AUD severity | | | | | |
| Fede, 2019 | MODERATE; 1 source but from community | MODERATE: N/S | LOW: yes | LOW: N/S | MODERATE |

| | | | | | |
|---|---|---|---|---|---|
| Risk factors for alcohol use | | | | | |
| Winham, 2016 | LOW; Multisite study | MODERATE: N/S | HIGH: no | LOW: N/S | MODERATE |
| Predicting future alcohol use | | | | | |
| King, 2011 | LOW; Large dataset across 6 countries | LOW | LOW: yes | LOW: N/S | LOW |
| Kinreich, 2019 | LOW; 9 sites across the US | MODERATE: N/S | HIGH: no | LOW: N/S | MODERATE |
| Prediction of Treatment Outcomes | | | | | |
| Symons, 2020 | HIGH; 1 site | HIGH | HIGH: no | LOW: N/S | HIGH |
| Symons, 2019 | HIGH; 1 site | HIGH | HIGH: no | LOW: N/S | HIGH |
| Lindner, 2020 | HIGH; 1 site | HIGH | HIGH: no | LOW: N/S | HIGH |
| Sekutowicz, 2019 | LOW; Multisite study | MODERATE: N/S | HIGH: no | LOW: N/S | MODERATE |
| Seo, 2015 | MODERATE; N/S | MODERATE: N/S | HIGH: no | LOW: N/S | HIGH |
| Winterer, 1998 A | HIGH; 1 site | MODERATE: N/S | MODERATE: N/S | LOW: N/S | HIGH |
| Winterer, 1998 B | HIGH; 1 site | MODERATE: N/S | MODERATE: N/S | LOW: N/S | MODERATE |
| Satapathy, 2020 | HIGH; 1 site | HIGH | HIGH: no | MODERATE: Patients with extensive history of prior rehabilitation did receive transplants | HIGH |
| Chih, 2014 | LOW; 2 sites | LOW | LOW: yes | LOW: N/S | LOW |
| Connor, 2007 | MODERATE; N/S | LOW | MODERATE: N/S | LOW: N/S | MODERATE |
| Kurth, 2001 | MODERATE; N/S | MODERATE: N/S | MODERATE: N/S | LOW: N/S | MODERATE |
| Hillemacher, 2012 | HIGH; 1 source | LOW | HIGH: no | LOW: N/S | HIGH |
| Burkhardt, 2020 | LOW; 2 sites | MODERATE | LOW: yes | LOW: N/S | MODERATE |
| Lapuerta, 1997 | MODERATE; N/S | MODERATE: N/S | LOW: yes | LOW: N/S | MODERATE |
| Choi, 2020 | LOW; Nationwide study Korea | MODERATE | LOW: yes | LOW: N/S | MODERATE |
| Hou, 2015 | LOW; 2 sites | LOW | HIGH: no | LOW: N/S | MODERATE |
| Wei, 2020 | LOW; 2 sites | LOW | MODERATE: N/S | LOW: N/S | MODERATE |
| Laska, 2020 | LOW; Multisite study | HIGH | MODERATE: N/S | LOW: N/S | MODERATE |
| Lin, 2020 | LOW; Multisite study | MODERATE | HIGH: no | HIGH: Cell heterogeneity from blood samples; Single population | HIGH |
| Predicting treatment seeking behavior | | | | | |
| Lee, 2019 | LOW; Multisite study | MODERATE | LOW: yes | LOW: N/S | MODERATE |

| AUD in Adolescents | | | | | |
|---|---|---|---|---|---|
| Squeglia, 2017 | LOW; Multisource study | LOW | HIGH: no | LOW: N/S | MODERATE |
| Whelan, 2014 | LOW; Multisite study | LOW | LOW: yes | LOW: N/S | LOW |
| Afzali, 2019 | LOW; 2 sites | LOW | HIGH: no | LOW: N/S | MODERATE |
| Ruan, 2019 | LOW; Multisite study | LOW | HIGH: no | LOW: N/S | MODERATE |
| Vázquez, 2020 | LOW; Multisource study | HIGH | MODERATE: N/S | LOW: N/S | MODERATE |
| García, 2009 | MODERATE; N/S | MODERATE: N/S | HIGH: no | LOW: N/S | HIGH |
| Weidacker, 2020 | LOW; Multisite study | MODERATE: N/S | HIGH: no | LOW: N/S | MODERATE |
| O'Halloran, 2020 | HIGH; 1 source | MODERATE: N/S | HIGH: no | LOW: N/S | HIGH |
| O'Halloran, 2018 | LOW; 2 sites | MODERATE: N/S | HIGH: no | LOW: N/S | MODERATE |
| Martinez-Loredo, 2018 | LOW; 22 sites | LOW | HIGH: no | LOW: N/S | MODERATE |

Abbreviations: Electroencephalographic (EEG); Electronic health record (EHR); Not specified (NS).
